# Gal-9 and DCN Serum Expression Reflect Accelerated Brain Aging and Are Attenuated by the Green-Mediterranean Diet: The 18-month DIRECT PLUS Proteomics-Brain MRI Trial

**DOI:** 10.1101/2024.11.19.24317485

**Authors:** Dafna Pachter, Anat Yaskolka Meir, Alon Kaplan, Gal Tsaban, Hila Zelicha, Ehud Rinott, Gidon Levakov, Ofek Finkelstein, Ilan Shelef, Moti Salti, Frauke Beyer, Veronica Witte, Nora Klöting, Berend Isermann, Uta Ceglarek, Tammy Riklin Raviv, Matthias Blüher, Michael Stumvoll, Dong D. Wang, Frank B Hu, Meir J Stampfer, Galia Avidan, Iris Shai

## Abstract

**Background:** We recently reported that a green-Mediterranean (green-MED), high-polyphenol diet is potentially neuroprotective for age-related brain atrophy. Here, we explored the interplay between dietary intervention, proteomics profile, and accelerated brain age.

**Methods:** In the 18-month DIRECT PLUS trial, 294 participants (adherence rate=89%) were randomized to one of three arms: 1) Healthy dietary guidelines (HDG); 2) MED diet; or 3) green-MED diet. Both MED diets included 28g/day of walnuts. Additionally, the low red/processed meat green-MED group received daily supplements of polyphenol-rich green-tea and green Mankai aquatic plant. In this secondary analysis, we measured 87 serum proteins (Olink-CVDII) and conducted Magnetic Resonance Imaging (MRI) to obtain brain 3D-T1-weighted for brain age calculation based on brain convolutional neural network to identify protein markers reflecting the brain age gap (BAG: residual deviation of MRI-assessed brain age from chronological age).

**Results:** We analyzed eligible brain MRIs (216 at baseline and 18-month) for BAG calculation. At baseline (age=51.3yrs, 90% men), lower weight, waist circumference, diastolic blood pressure, and HbA1c parameters were associated with younger brain age than expected (p<0.05 for all). At baseline, higher levels of two specific proteins: Galectin-9 (Gal-9) and Decorin (DCN), were associated with larger BAG (accelerated brain aging; FDR<0.05). A proteomics principal component analysis (PCA) revealed a significant difference between the 18-month time points among participants who completed the trial with accelerated brain aging (p=0.02). Between baseline and 18 months, Gal-9 significantly decreased (p<0.05) among individuals who completed the intervention with attenuated brain age, and DCN significantly increased (p<0.05) among those who completed the trial with accelerated brain age. A significant interaction was observed between the green-MED diet and proteomics PCA change compared to the HDG (β=-1.7; p-interaction=0.05). Participants in the green-MED diet significantly decreased Gal-9 compared to the HDG diet (p=0.015) and from baseline (p=0.003). DCN levels, however, marginally increased in the HDG diet from baseline (p=0.053).

**Conclusion:** Higher serum levels of Gal-9 and DCN may indicate an acceleration of brain aging and might be reduced by the green-MED/high-polyphenol diet rich in Mankai and green-tea and low in red/processed meat.

**Trial registration number:** NCT03020186.

## Introduction

Age-related neurodegenerative conditions are often characterized by reduced brain volume (atrophy), as well as enlargement of ventricles and cerebral-spinal fluid (CSF) spaces (1). However, these changes do not always correspond with chronological age. Factors such as diabetes, inflammation, hypertension, high cholesterol, and accumulation of β amyloid and tau markers can accelerate brain aging (2–10).

In recent years, brain age has emerged as a promising index of overall brain health (10–13). Brain age gap (BAG) is the difference between an individual’s brain age based on MRI and their chronological age. A positive BAG reflects brain age higher than chronological age or accelerated brain aging. In contrast, a negative BAG indicates brain age lower than chronological age or attenuated brain aging (14). Higher brain age, in relation to chronological age, is observed in various neurological conditions, including mild cognitive impairment (MCI) and Alzheimer’s disease (AD). However, no studies of circulating proteomics as an indicator of BAG have been published.

Recently, in the 18-month DIRECT PLUS trial, we found that a green-MED (high-polyphenol) diet, rich in Mankai, green tea, and walnuts and low in red/processed meat, is potentially neuroprotective for age-related brain atrophy (15). We used data from this trial to identify proteomic signatures reflecting BAG status at baseline and at 18 months, and the impact of lifestyle interventions on those proteomics signatures.

## Methods

### Study Recruitment

The 18-month DIRECT PLUS trial included 294 participants and was conducted in an isolated workplace (Nuclear Research Center Negev, Dimona, Israel). The inclusion criteria were age ≥30 years with abdominal obesity (waist circumference (WC): men>102 cm, women >88 cm) or dyslipidemia (triglycerides>150 mg/dL; high-density lipoprotein cholesterol (HDLc) ≤40mg/dL for men, ≤50mg/dL for women). (Exclusion criteria are detailed in **Supplementary Methods 1**). The study was approved by the Institutional Review Board at Soroka University Medical Center. The trial was conducted in a single phase from May 2017 to November 2018. Participants provided written informed consent and did not receive compensation for their participation.

### Study design, randomization, and intervention

Participants were randomly assigned in a 1:1:1 ratio to one of three intervention groups: 1) HDG, an active control group; 2) a traditional calorie-restricted Mediterranean (MED) diet, low in simple carbohydrates; or 3) the green-MED diet. Randomization was conducted in a single phase using a parallel assignment model, and participants were aware of their assigned intervention (open-label protocol). Each group received specific nutritional guidance along with physical activity (PA) instructions. The HDG group received basic health-promoting dietary guidelines. The MED group received guidelines for a calorie-restricted traditional MED diet, low in simple carbohydrates, as described in our previous papers (16, 17).

The MED diet was rich in vegetables, with beef and lamb replaced by poultry and fish. Both MED diets included 28 grams of walnuts per day. The green-MED diet, in addition to 28 grams of walnuts per day, was lower in processed and red meat compared to the MED diet and richer in plants and polyphenols. This was achieved through the consumption of 3-4 cups of green tea and 500 ml of a Mankai-based (cultivated duckweed) product daily (18–20), a green shake at dinner. Both MED diets were equally calorie-restricted, with 1500–1800 kcal/day for males and 1200–1400 kcal/day for females. All participants were provided with a free gym membership and PA guidelines. Additionally, they attended periodic 90-minute nutritional and PA sessions at their workplace, facilitated by a multidisciplinary team of physicians, clinical dietitians, and fitness instructors.

### Clinical measurements

Clinical and anthropometric biomarkers were measured at baseline and after 18 months. Height was measured to the nearest millimeter using a standard wall-mounted stadiometer. Body weight was measured without shoes to the nearest 0.1 kg. Waist circumference (WC) was measured halfway between the lowest rib and the iliac crest to the nearest millimeter using standard procedures and an anthropometric measuring tape. Two blood pressure (BP) and heart rate measurements were recorded after resting using an automatic BP monitor, and BP was calculated as the mean of the two measurements. Blood samples were taken at 8 am after a 12-hour fast, and aliquots were frozen for later assays. (Further laboratory methods are detailed in **Supplementary Methods 2**).

### Proteomic panel

We assayed available serum samples for proteomic analysis from baseline (N=211) and 18-months (N=208), focusing on participants with complete data at baseline and after the 18-month intervention. The protein analysis was conducted on 92 proteins with Olink’s CARDIOVASCULAR II panel on their proteomics platform. Five proteins with more than 5% missing data (BNP, ITGB1BP2, PARP1, SERPINA12, STK4) were excluded. The proteomics platform uses proximity extension assay (PEA) technology in a 96-well plate format. Each panel contains 92 pairs of oligonucleotide-labeled antibody probes that attach to their target proteins in serum. Following a proximity-dependent DNA polymerization event, a PCR reporter sequence is created, amplified, and measured using real-time PCR. Internal and external controls are employed for data normalization and quality assurance. Intra- and inter-coefficient variance (CV)% were calculated from control samples (pooled plasma samples) included on each plate. This platform provides data on normalized protein expression (NPX) using a log2 scale. More details about the Olink analysis are available in **Supplementary Methods 3**.

### Brain MRI acquisition and brain age prediction

We analyzed eligible brain MRIs (216 at baseline and 18-m) for BAG calculation. (**Supplementary Figure 1**). MRI scans were conducted at the Soroka University Medical Center, Beer Sheva using a 3 T Philips Ingenia scanner (Amsterdam, The Netherlands) equipped with a standard head coil. High-resolution anatomical volumes were obtained using a T1-weighted 3D pulse sequence (1 × 1 × 1 mm3, 150 slices, TR = 2500, TE = 30 ms, field of view 240 × 220 × 150). The brain age was predicted based on a previously established convolutional neural network (CNN) model using a sample of 19,275 brain images from various open datasets including different MRI scanners, randomly divided into training (N = 13,471; 70%), validation (N = 1957; 10%), and testing (N = 3847; 20%) (21). The fitted model was then applied to the 216 participants from the DIRECT PLUS study. According to previous findings (22), the model generalized better on the test set, with an MAE of 2.96 years and Pearson correlation of 0.94. For the DIRECT PLUS dataset, the model performed similarly at baseline time-point and 18-month time-point, with MAEs of 5.03 and 4.96 years and Pearson correlations of 0.82 and 0.83, respectively (21). The BAG was determined by calculating the difference in years between the predicted brain age and the chronological age.

### Statistical analysis

BAG calculations at baseline and 18 months were conducted using the residuals from the linear regression between brain age and chronological age. We stratified baseline characteristics by positive (accelerated brain aging) or negative (attenuated brain aging) BAG. Continuous variables are presented as mean ± SD and median ± IQR depending on their distribution and as number (%) for categorical variables. Statistical tests used include Welch Two Sample t-test, Pearson’s Chi-squared test, and Wilcoxon rank sum test. Spearman correlations were calculated to quantify the associations between proteomics levels and BAG. The false discovery rate (FDR<0.05) was applied to correct for multiple testing. Partial correlations were calculated to adjust for age and sex. Elastic net regression with leave-one-out cross-validation (LOOCV) was used to establish proteomic scores to predict the BAG. Baseline data were used as training and test sets, and 18-month post-intervention data were used for validation.

For the 18-month change analyses, we examined differences in the proteomics panel change between baseline and 18-month time points using principal component analysis (PCA). Linear Mixed Effects Models (LMM) were conducted to examine the difference at baseline and 18 months for those completed with positive or negative BAG and BAG change. In addition, a volcano plot was applied to phenotype the proteomics for each brain age gap group. Further, we conducted a quantile regression of chronological age for candidate proteins. In addition, we used LMM to test for the interaction between the lifestyle intervention groups and the two-time points to predict the change in PC1 for the full proteomics. Furthermore, we quantified the change in levels of specific proteins for each lifestyle intervention group by ANOVA after Bonferroni correction. LMM was used to compare the 18-month changes in protein markers from baseline, also adjusted for Bonferroni comparisons. Statistical analysis was performed using R (Version 4.2.0), details regarding the specific R packages used are provided in **Supplementary Methods 4**. Statistical significance was set at a two-sided alpha value of 0.05.

## Results

### Baseline characteristics of the participants

Chronological age and brain age were highly correlated (r=0.82, p>0.001; **Figure 1**).

**Figure 1:**
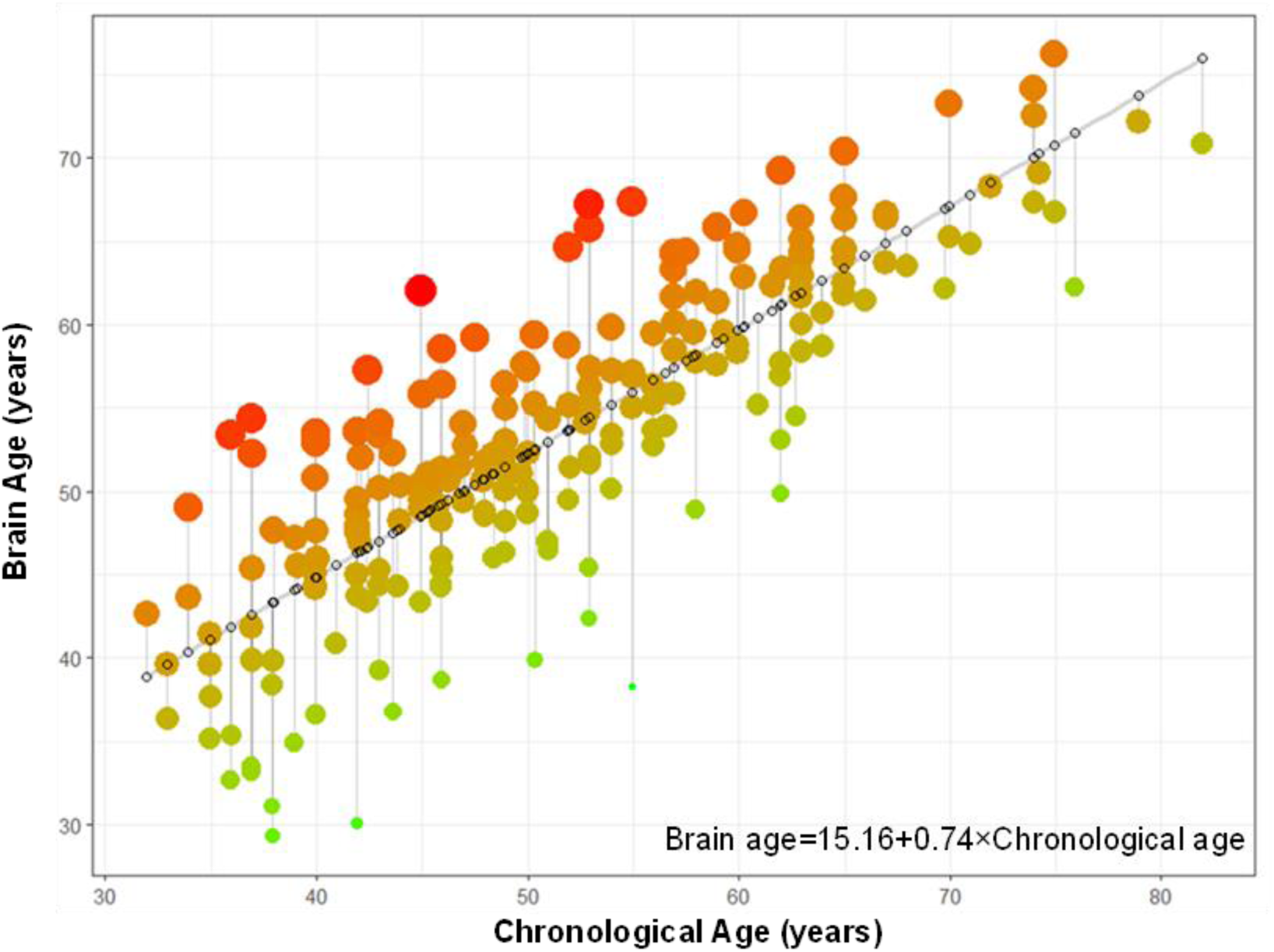
Correlation of chronological age and brain age and Brain Age Gap representation. N=216. Brain Age Gap (BAG) was determined by linear regression with brain age as the dependent variable and chronological age as the independent variable. BAG < 0 represents delayed aging (expected brain age > observed brain age); BAG > 0 represents accelerated aging (observed brain age > expected brain age). The color and size of the points indicate the magnitude of the residuals, with green and smaller dots showing smaller residuals and red and larger dots showing larger residuals.

Baseline characteristics of the 216 participants stratified by positive (accelerated brain aging-older brain than expected for their chronological age) and negative (attenuated brain aging-younger brain than expected for their chronological age) BAG are presented in **Table 1**. The mean BAG of the positive group (accelerated brain aging) was 4.0 ± 3.2 years, and for the negative group (attenuated brain aging), −4.2 ± 3.8 years. Participants with younger brain age had a lower weight, WC, diastolic BP, and HbA1c (p<0.05 for all).

**Table 1:**
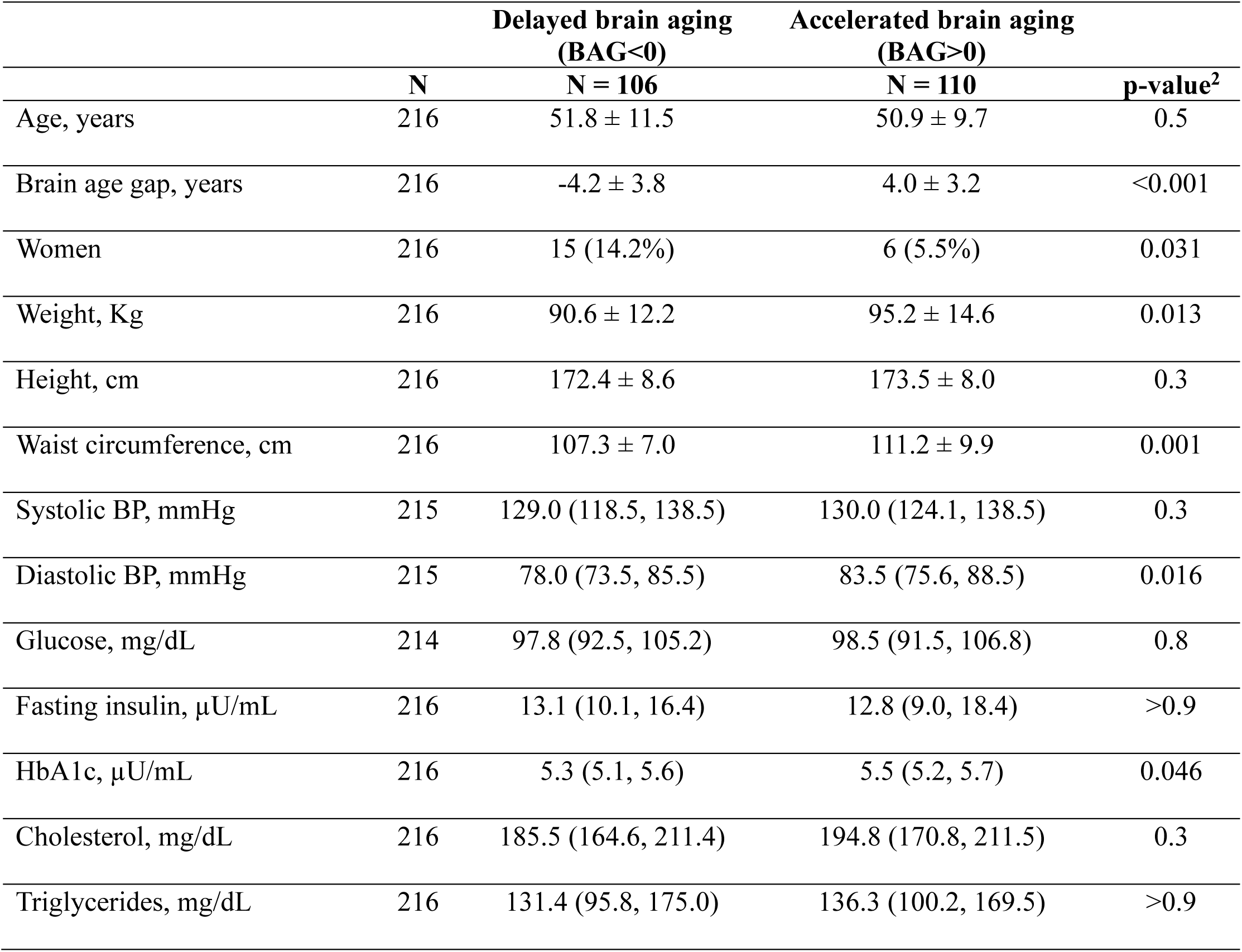
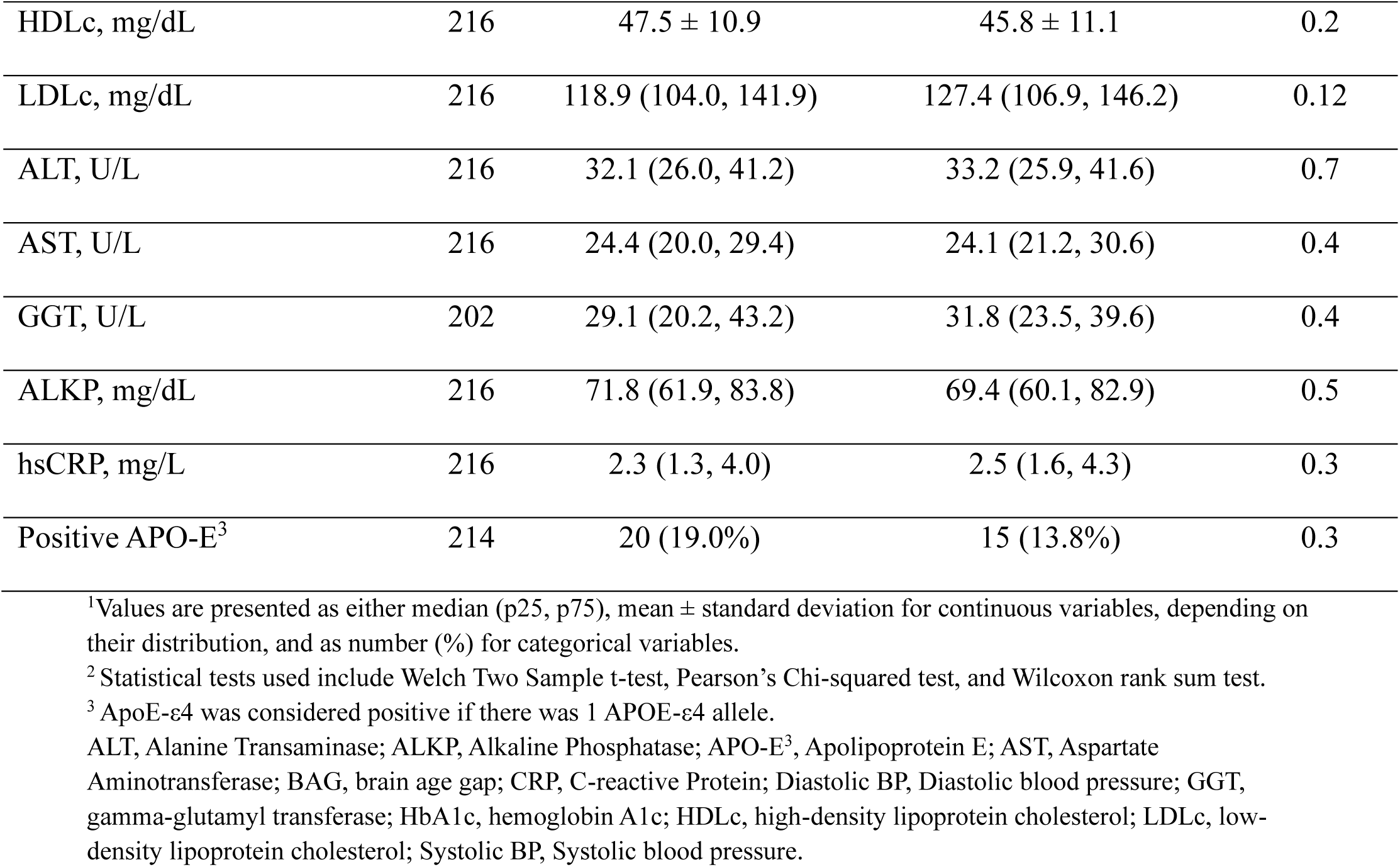
Baseline characteristics of the DIRECT PLUS participants stratified by delayed/accelerated brain aging^1^.

|  |  | Delayed brain aging<br>(BAG<0) | Accelerated brain aging<br>(BAG>0) |  |
| --- | --- | --- | --- | --- |
|  | N | N = 106 | N = 110 | p-value<sup>2</sup> |
| Age, years | 216 | $51.8 \pm 11.5$ | $50.9 \pm 9.7$ | 0.5 |
| Brain age gap, years | 216 | $-4.2 \pm 3.8$ | $4.0 \pm 3.2$ | <0.001 |
| Women | 216 | 15 (14.2%) | 6 (5.5%) | 0.031 |
| Weight, Kg | 216 | $90.6 \pm 12.2$ | $95.2 \pm 14.6$ | 0.013 |
| Height, cm | 216 | $172.4 \pm 8.6$ | $173.5 \pm 8.0$ | 0.3 |
| Waist circumference, cm | 216 | $107.3 \pm 7.0$ | $111.2 \pm 9.9$ | 0.001 |
| Systolic BP, mmHg | 215 | 129.0 (118.5, 138.5) | 130.0 (124.1, 138.5) | 0.3 |
| Diastolic BP, mmHg | 215 | 78.0 (73.5, 85.5) | 83.5 (75.6, 88.5) | 0.016 |
| Glucose, mg/dL | 214 | 97.8 (92.5, 105.2) | 98.5 (91.5, 106.8) | 0.8 |
| Fasting insulin, μU/mL | 216 | 13.1 (10.1, 16.4) | 12.8 (9.0, 18.4) | >0.9 |
| HbA1c, μU/mL | 216 | 5.3 (5.1, 5.6) | 5.5 (5.2, 5.7) | 0.046 |
| Cholesterol, mg/dL | 216 | 185.5 (164.6, 211.4) | 194.8 (170.8, 211.5) | 0.3 |
| Triglycerides, mg/dL | 216 | 131.4 (95.8, 175.0) | 136.3 (100.2, 169.5) | >0.9 |
| HDLc, mg/dL | 216 | 47.5 ± 10.9 | 45.8 ± 11.1 | 0.2 |
| LDLc, mg/dL | 216 | 118.9 (104.0, 141.9) | 127.4 (106.9, 146.2) | 0.12 |
| ALT, U/L | 216 | 32.1 (26.0, 41.2) | 33.2 (25.9, 41.6) | 0.7 |
| AST, U/L | 216 | 24.4 (20.0, 29.4) | 24.1 (21.2, 30.6) | 0.4 |
| GGT, U/L | 202 | 29.1 (20.2, 43.2) | 31.8 (23.5, 39.6) | 0.4 |
| ALKP, mg/dL | 216 | 71.8 (61.9, 83.8) | 69.4 (60.1, 82.9) | 0.5 |
| hsCRP, mg/L | 216 | 2.3 (1.3, 4.0) | 2.5 (1.6, 4.3) | 0.3 |
| Positive APO-E<sup>3</sup> | 214 | 20 (19.0%) | 15 (13.8%) | 0.3 |

<sup>1</sup>Values are presented as either median (p25, p75), mean ± standard deviation for continuous variables, depending on their distribution, and as number (%) for categorical variables.
<sup>2</sup>Statistical tests used include Welch Two Sample t-test, Pearson's Chi-squared test, and Wilcoxon rank sum test.
<sup>3</sup>ApoE-ε4 was considered positive if there was 1 APOE-ε4 allele.
ALT, Alanine Transaminase; ALKP, Alkaline Phosphatase; APO-E<sup>3</sup>, Apolipoprotein E; AST, Aspartate Aminotransferase; BAG, brain age gap; CRP, C-reactive Protein; Diastolic BP, Diastolic blood pressure; GGT, gamma-glutamyl transferase; HbA1c, hemoglobin A1c; HDLc, high-density lipoprotein cholesterol; LDLc, low-density lipoprotein cholesterol; Systolic BP, Systolic blood pressure.

### Cross-sectional associations between proteomics and Brain Age Gap at baseline

Baseline correlation analysis between BAG and 87 proteins of the proteomic panel revealed only two protein markers, Galectin 9 (Gal-9) and Decorin (DCN), both of which were significantly positively associated with greater BAG (FDR< 0.05; sex and age-adjusted) (Of the 216 participants with BAG, we had proteomics available for 211; **Figure 2**).

**Figure 2:**
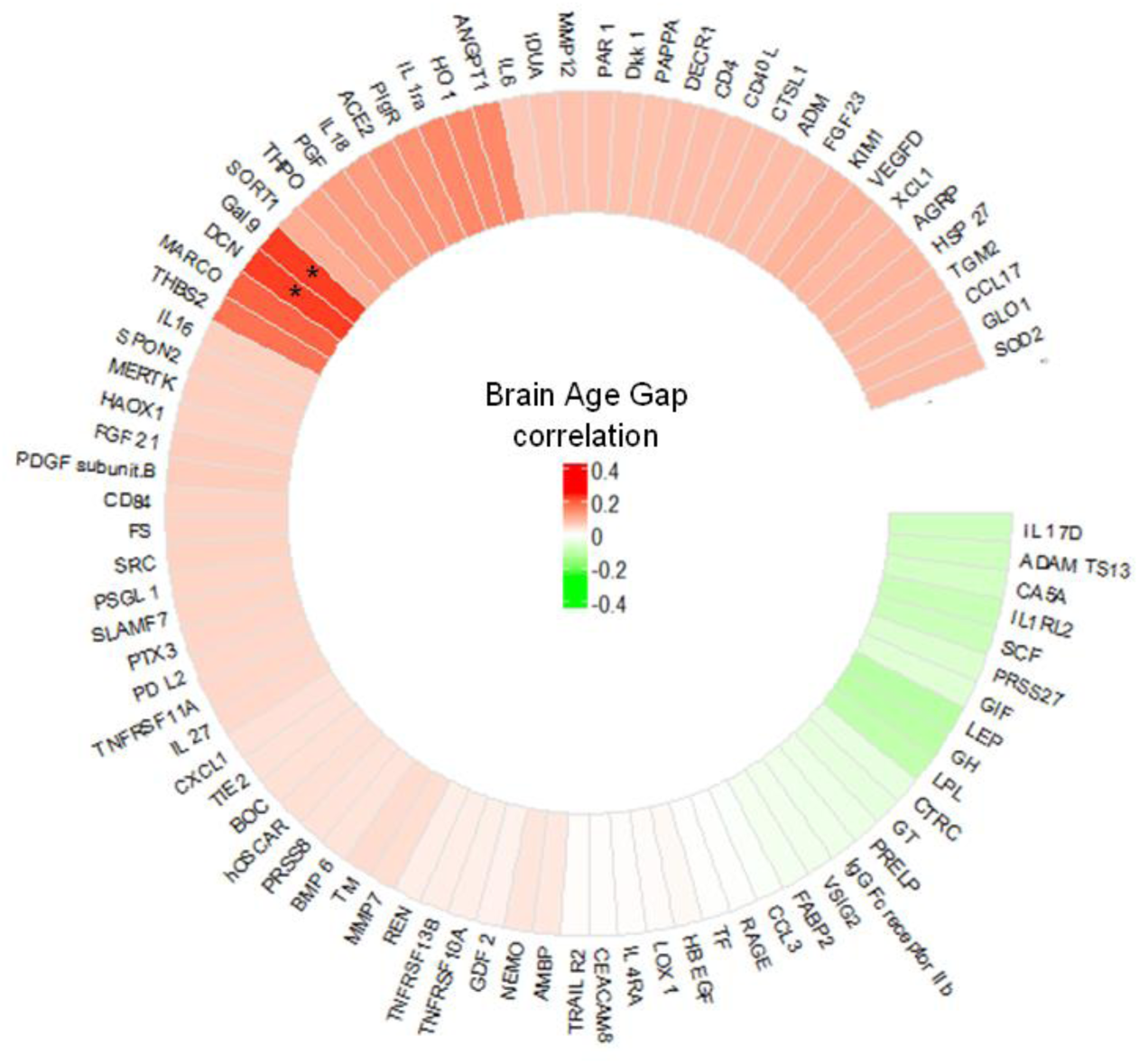
Baseline correlation between Brain Age Gap and proteomic panel. Circular heatmap of the proteomic panel (N=87 proteins). Spearman correlation test was used to test the correlations between baseline Brain Age Gap and each protein. Correlations adjusted for sex and age. * Denotes significant correlation at FDR < 0.05 (N=211).

### Proteomic score for Brain Age Gap

Three models were utilized to determine whether proteomics significantly adds to other known or suspected markers to improve the prediction accuracy of BAG (**Table 2**, **Supplementary Figure 2**). The first model (model i) only included the proteomic panel at baseline (87 proteins) and had an R squared (R^2^) of 0.034 and root mean square error (RMSE) of 5.34. The 5 predictors of model i, according to their importance, were Gal-9, DCN, ANGPT1, THBS2, and LEP (**Supplementary Figure 2A**). The second model (model ii) included only the traditional biomarkers at baseline (16 biomarkers: WC, BMI, systolic BP, diastolic BP, glucose, HbA1c, insulin, cholesterol, triglycerides, high-density lipoprotein cholesterol (HDLc), low-density lipoprotein cholesterol (LDLc), alkaline phosphatase (ALKP), alanine transaminase (ALT), aspartate aminotransferase (AST), gamma-glutamyl transferase (GGT), c-reactive protein (CRP)) had an R^2^ of 0.003 and RMSE of 67.75. The 6 predictors of model ii, according to importance, were WC, AST, SBP, DBP, ALKP, and HDLc (**Supplementary Figure 2B**). The third model (model iii) tests whether a combination of the proteomic panel with traditional biomarkers would improve the prediction accuracy of BAG. This model yielded an R^2^ of 0.041 and an RMSE of 5.16. The 10 first predictors of model 3, according to the importance, were Gal-9, DCN, MARCO, VEGFD, WC, PIgR, DBP, SBP, AST, ALKP, HDLc, ADAM-TS13, LEP (**Supplementary Figure 2C**). Next, we validated the models presented in **Table 2** on an 18-month time point from our DIRECT PLUS trial. Pearson R values for the validation sets were (model i, n=210: r=0.263; p=1.09e-04, model ii, n=213: r=0.275; p= p=4.76e-05, model iii, n=210: r=0.372; p=3.52e-08, **Supplementary Table 1**).

**Table 2:**
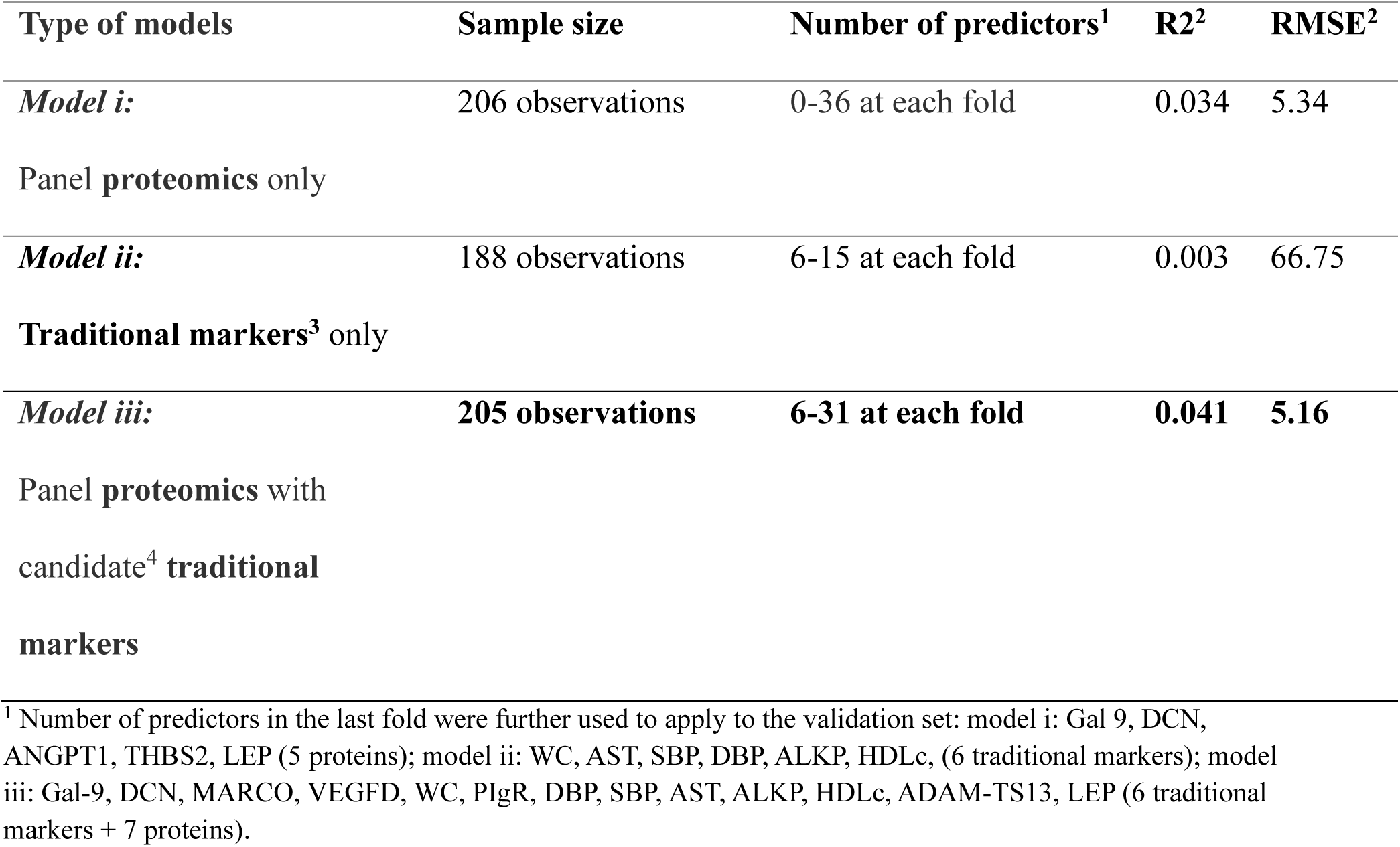
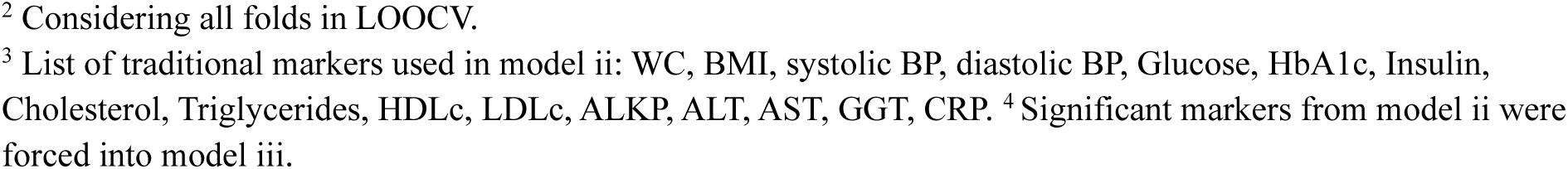
Brain Age Gap prediction by the proteomic panel and traditional variables.

| Type of models | Sample size | Number of predictors<sup>1</sup> | R<sup>2</sup> | RMSE<sup>2</sup> |
| --- | --- | --- | --- | --- |
| *Model i:* | 206 observations | 0-36 at each fold | 0.034 | 5.34 |
| Panel **proteomics** only |  |  |  |  |
| *Model ii:* | 188 observations | 6-15 at each fold | 0.003 | 66.75 |
| **Traditional markers<sup>3</sup>** only |  |  |  |  |
| *Model iii:* | **205 observations** | **6-31 at each fold** | **0.041** | **5.16** |
| Panel **proteomics** with<br>candidate<sup>4</sup> **traditional**<br>**markers** |  |  |  |  |

<sup>1</sup> Number of predictors in the last fold were further used to apply to the validation set: model i: Gal 9, DCN, ANGPT1, THBS2, LEP (5 proteins); model ii: WC, AST, SBP, DBP, ALKP, HDLc, (6 traditional markers); model iii: Gal-9, DCN, MARCO, VEGFD, WC, PIgR, DBP, SBP, AST, ALKP, HDLc, ADAM-TS13, LEP (6 traditional markers + 7 proteins).
<sup>2</sup> Considering all folds in LOOCV.
<sup>3</sup> List of traditional markers used in model ii: WC, BMI, systolic BP, diastolic BP, Glucose, HbA1c, Insulin, Cholesterol, Triglycerides, HDLc, LDLc, ALKP, ALT, AST, GGT, CRP. <sup>4</sup> Significant markers from model ii were forced into model iii.

### Post-intervention distinct Brain Age Gap groups and their proteomic phenotype

Next, we examined differences in the proteomics panel change between baseline and 18 months using PCA analysis for proteomics. Of the 216 participants with BAG, we had proteomics available for 208 at both time points; **Figure 3A**. Using LMM, we found that the first principal component (PC1) was significantly different between the two-time points (p=0.03). Further, we divided participants into two distinct groups of BAG change: participants who completed the 18-month intervention with negative BAG (younger brain than expected, N=102) and participants who completed the intervention with positive BAG (older brain than expected, N=106) (**Figure 3B-C**). The PCA analysis for each group between the two-time points showed that PC1 was significantly different between the two-time point groups in participants who completed the intervention with an older brain than expected (p=0.02). However, the difference was not significant in those who completed the intervention with a younger brain than expected (p=0.5).

**Figure 3:**
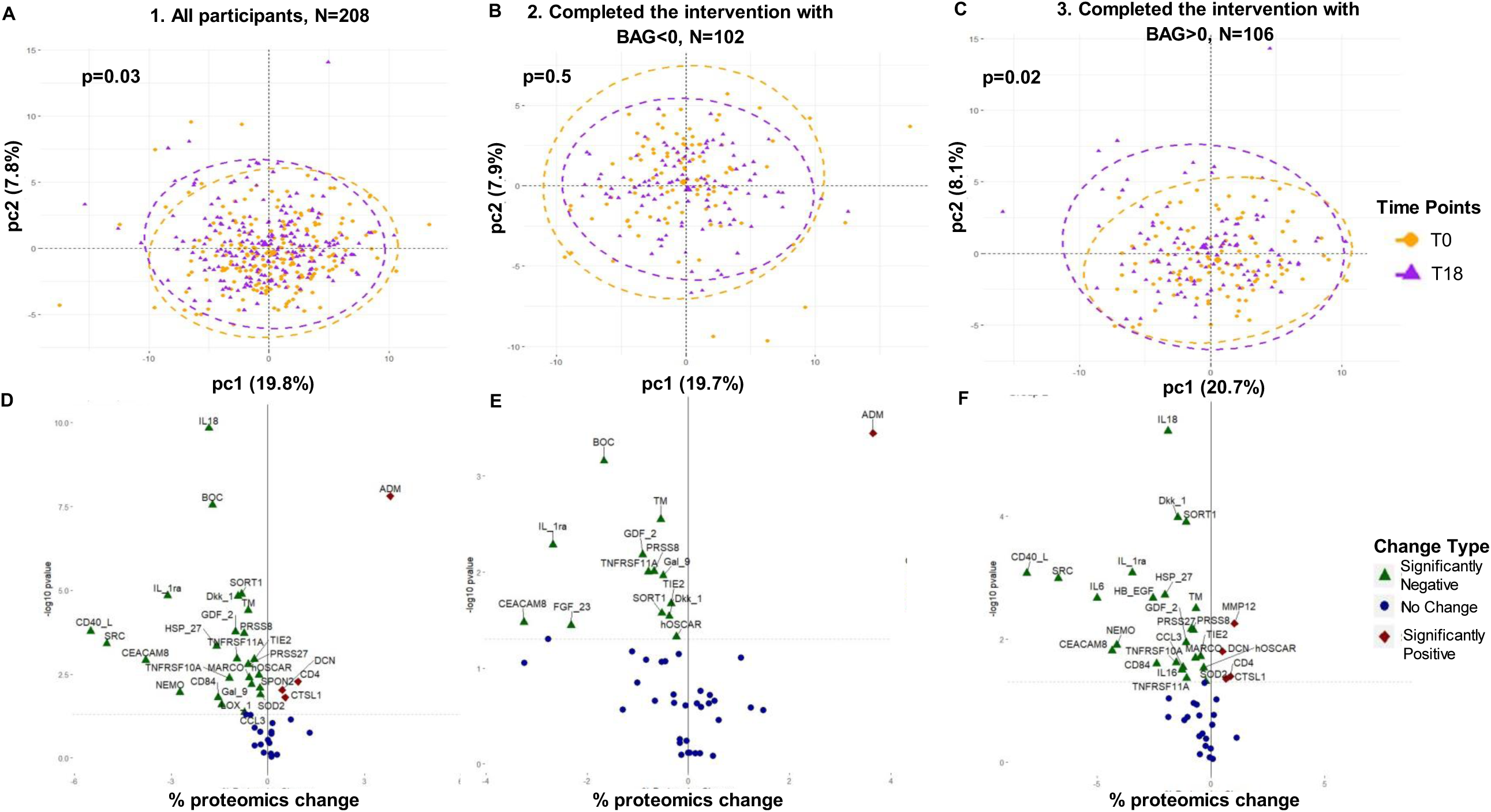
Distinct Brain Age Gap switch groups and their proteomic phenotype. Panel 1. All participants, n=208. Panel 2. Participants who completed with BAG<0, n=102 (T0 + T18 with BAG <0 or T0 BAG >0 & T18 BAG<0). Panel 3. Participants who completed with BAG>0, n=106 (T0 + T18 with BAG >0 or T0 BAG <0 & T18 BAG >0). PCA plots (A, B, C): PCA analysis for proteomics in distinct panel sub-groups, including baseline (T0) and 18-month time-points (T18). PC1 and PC2 are presented. P values for LMM between time points of PC1 are presented. Volcano plots (D, E, F): % proteomics change which correlated with PC1 (absolute correlation value > 0.1) was calculated relative to the baseline protein’s expression. The statistical significance of the proteomics change (T18/T0) was analyzed using the Wilcoxon signed-rank test. The rhombus shape represents significant proteomics with a negative % change. The triangular shape represents significant proteomics with a positive % change. The circle shape represents non-significant proteomics.

We further explored the individual proteins that most strongly predicted BAG change, with an absolute correlation higher than 0.1, for BAG change (for the proteomics panel list for each group, please see **Supplementary Table 2**). For all participants, we found significant positive and negative relative changes for DCN and Gal-9, respectively (Wilcoxon signed-rank test is applied, p<0.05) (**Figure 3D; Supplementary Table 3**). Our findings aligned with our initial results, a negative Gal-9 relative change to the baseline protein levels in participants who completed with a younger brain than expected and a positive DCN relative change to the baseline protein levels in participants who completed with a younger brain than expected (**Figure 3E-F; Supplementary Table 3**; p<0.05 for both).

### Dynamics in Gal-9 and DCN levels in aging and diet groups

In the subsequent analysis, we examine whether specific age ranges exhibit a different level of DCN and Gal-9 markers, potential brain aging indicators. To this end, we conducted a quantile regression, dividing chronological age into quantiles and testing whether any quantile group displayed a significant correlation between age and DCN or Gal-9 markers (**Figure 4A-B**).

**Figure 4:**
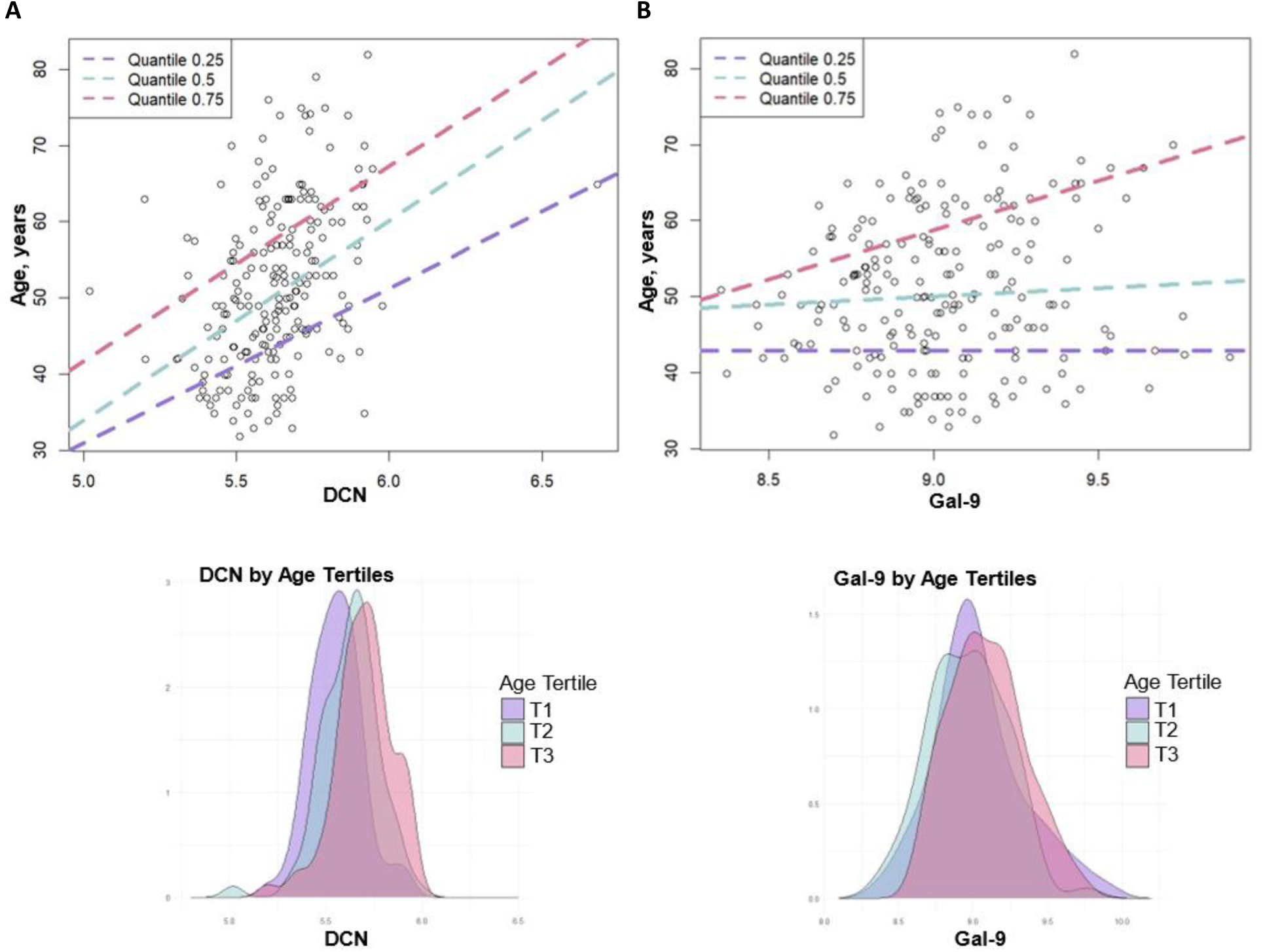
Chronological age by the candidate proteins: DCN and Gal 9. Scatter plots of quantile regression at baseline and density plots of the candidate proteins by tertiles of chronological age. A. DCN; B. Gal-9.

We found a statistically significant association in the Gal-9 marker only among individuals of the highest age group (tau= 0.75, coefficients= 12.98, p<0.001, mean age tertile = 63.7 years). Whereas, for DCN, a significant association was evident across all age groups, with a notably higher coefficient observed within the middle and higher quantiles (tau= 0.25, coefficients= 20.21, mean age tertile = 40.1 years, tau= 0.50, coefficients= 26.17, mean age tertile = 50.2 years, tau= 0.75, coefficients= 25.58, mean age tertile = 63.7 years, p<0.001 for all; **Table 3**).

**Table 3:** Quantile regression for prediction of chronological age by DCN and Gal-9.

| Proteins | Y=Age at baseline | Tau=0.25 | Tau=0.50 | Tau=0.75 |
| --- | --- | --- | --- | --- |
| **Gal-9** |  |  |  |  |
|  | Intercept | 42.92 | 30.08 | -58.00 |
|  | B | 0 | 2.11 | **12.98<sup>1</sup>** |
| **DCN** |  |  |  |  |
|  | Intercept | -69.98 | -96.89 | -86.21 |
|  | B | **20.21<sup>1</sup>** | **26.17<sup>1</sup>** | **25.58<sup>1</sup>** |
|  | **Mean Tertile Age (Years)<sup>2</sup>** | 40.1 | 50.2 | 63.7 |

<sup>1</sup> Denotes p<0.05
<sup>2</sup> Mean tertile groups (T1, T2, T3), density plots represent the Gal-9/DCN distribution across these tertiles.

Using LMM, we also tested the interaction between the lifestyle intervention groups and the two-time points for the association with PC1. We found that the proteomics in the green-MED group showed a significant change during the intervention (β = −1.7; p of interaction = 0.05) compared to the HDG group. Finally, we quantified the specific change in DCN and Gal-9 biomarkers for each lifestyle intervention group. For Gal-9, the Green-MED group experienced a mean± se change of −0.104 ± 0.03, whereas Gal-9 changed only slightly in the MED (−0.03 ± 0.03) and HDG (0.002 ± 0.03) groups. Participants in the green-MED diet had a significant reduction in Gal-9 compared to the HDG diet after Bonferroni correction (p=0.015), and only the green-MED had a significant reduction compared to the baseline (p=0.003). Whereas green-MED, MED, and HDG diets represent the mean± se DCN change of 0.008 ± 0.02, 0.039 ± 0.02, and 0.032 ± 0.013, respectively, with no significant difference between diets. In the HDG group, DCN tended to increase following the intervention compared to the baseline (p = 0.053) (**Figure 5**).

**Figure 5:**
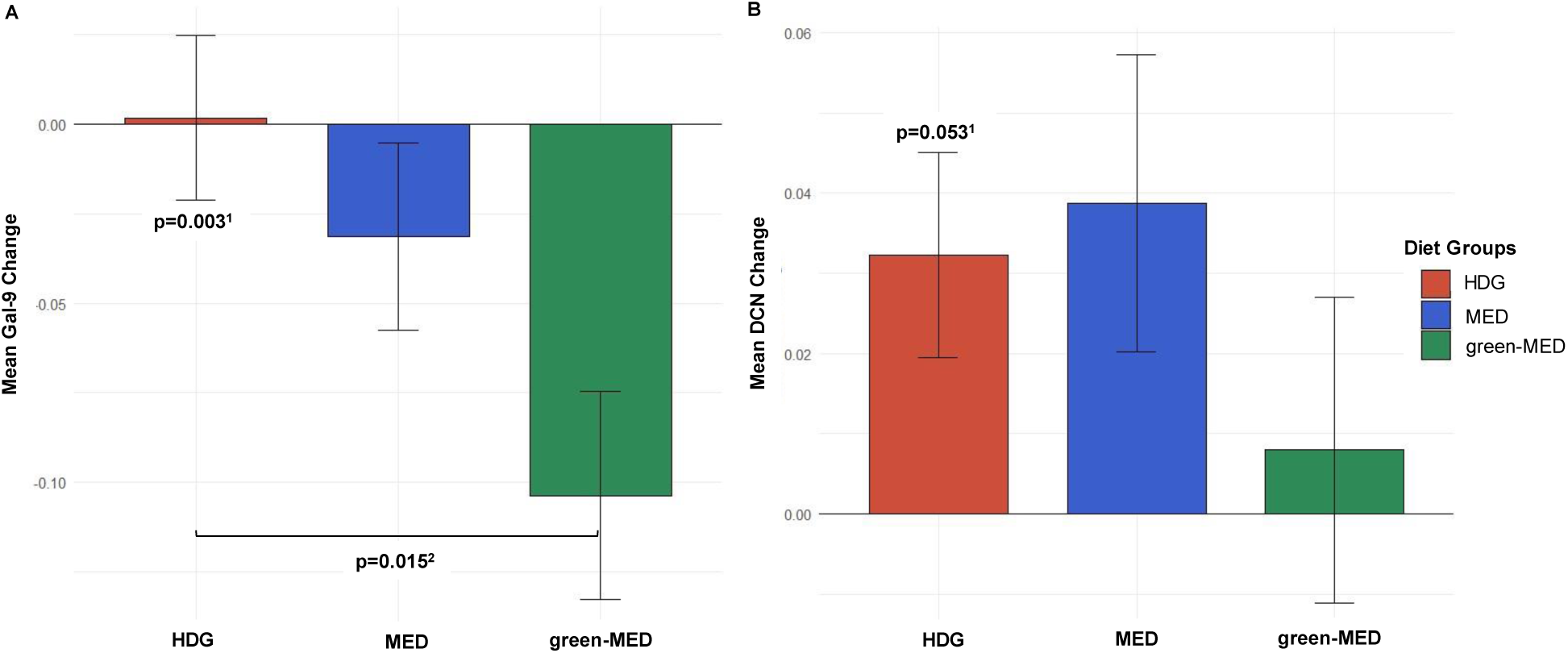
18-month changes in Gal-9 and DCN stratified by diet intervention groups. A: The Gal-9 mean ± SE changes are 0.002± 0.03 for HDG (n=70), −0.03± 0.03 for Med (n=70), and −0.104 ± 0.03 for Green Med (n=76), respectively. B: The DCN mean ± SE changes are 0.008 ± 0.02, 0.039 ± 0.02, and 0.032 ± 0.013 for green-MED (n=76), MED (n=70), and HDG (n=70), respectively. The mean and standard error are displayed on the graph for each group. “1”-p-value by LMM between time 0 and time 18 after Bonferroni correction. “2”-p-value between groups after Bonferroni correction.

## Discussion

We found two specific serum protein expressions, Gal-9 and DCN, that reflect accelerated brain aging. These proteomic markers can improve the prediction of BAG together with traditional variables. We further showed that levels of Gal-9 and DCN might favorably be modified by the green-MED/ high-polyphenol diet and that these changes are associated with better BAG. These findings are closely aligned with our previous results shadowing that the green-MED, high-polyphenol diet, rich in Mankai, green tea, and walnuts and low in red/processed meat, is potentially neuroprotective for age-related brain atrophy (15).

Several limitations need to be acknowledged. First, the high proportion of male participants (90%) may limit the generalizability of the findings to females. Additionally, the generalizability is further restricted since the participants had either abdominal obesity or dyslipidemia. Moreover, our proteomic assessment was limited to 87 proteins. In addition, this study lacks data about the participants’ educational or cognitive status. Indeed, in contrast to the significant brain atrophy and MRI-based BAG changes by diet group, we previously reported that we could not detect significant cognitive function differences between the diet groups following the intervention, possibly due to the small sample size of the sub-study, the limited sensitivity of the cognitive decline assessment methods or the short intervention time (15). However, the objectively measured BAG indicates early signs of cognitive decline and Alzheimer’s (23–25). Also, in the elastic net analysis for model ii, we encountered 0 predictors across multiple folds during the training step. However, we intentionally used a rigorous model with BAG value as the dependent variable rather than just the brain age to remove the effect of chronological age. Finally, we did not conduct an external validation of the elastic net model for BAG prediction. Nonetheless, internal validation sets at time 18 showed significant results (**Supplementary Table 1**). The strengths of this study include a relatively large sample size, a high retention rate, long-term intervention, and using the gold-standard brain MRI measurements for brain age calculation (21, 22).

In 1986, DCN was discovered to be a structural constituent of the extracellular matrix (ECM) (26). Since then, understanding of its role in various biological processes, including cell growth, differentiation, proliferation, and the regulation of inflammation and fibrillogenesis (27–29). Higher levels of DCN in cerebrospinal fluid (CSF) are associated with early Aβ pathology. Increased CSF-DCN levels are present at an early stage of amyloid-β (Aβ) amyloidosis and correlate with core AD biomarkers such as CSF-Aβ42 and CSF-p-tau. Additionally, DCN has been found to activate neuronal autophagy by enhancing autophagosomal-lysosomal degradation, linking ECM changes to autophagy (30, 31). Moreover, DCN pathway is related to learning and recall in mice (32).

Gal-9 is a member of the family of galactoside-binding proteins. Among the 15 galectin family members, galectins 1, 3, 4, 8, and 9 are significantly expressed in the brain and play crucial roles in neuromodulation, neuroinflammation, and neuroprotection (33). Gal-9 has been implicated in the modulation of immune responses as it induces the release of proinflammatory cytokines, initiating an inflammatory cascade (34, 35). Moreover, recombinant Gal-9 suppresses T lymphocytes but induces oligodendrocyte maturation and myelin repair in mixed glial cultures (36). Also, Gal-9 is expressed in astrocytes of multiple sclerotic lesions in the CSF of patients with multiple sclerosis (37). Furthermore, both ileal bile acid binding protein (I-BABP) and Gal-9 levels were elevated in AD patients (38). Similarly, Gal-3, another member of the galectin family (not included in our proteomic panel), is expressed in microglia clustered around Aβ plaques particularly in the hippocampus compared to the frontal cortex in AD patients. Higher Gal-3 levels are correlated with tau and p-Tau181 levels, two indicators of pathology progression in AD (39, 40).

Participants in the green-MED diet had a reduction in Gal-9 as compared to the HDG, and only the green-MED group had a significant reduction compared to the baseline. DCN increased in all diet groups, but least (not statistically significant) in the green-MED group. We previously reported that the green-MED (high-polyphenol) diet, rich in Mankai, green tea, walnuts, and low in red/processed meat, is potentially neuroprotective for age-related brain atrophy (15). The polyphenols in this diet may play a role in neuroprotection by reducing the activation of microglia, which is responsible for the inflammatory response in the central nervous system (41). As mentioned, DCN takes part in the regulation of inflammation and fibrillogenesis, and Gal-9 induces the release of proinflammatory. These mechanisms may explain the potential role of green-MED (high-polyphenol) in the reduction of the levels of these proteins and improving brain age.

In conclusion, BAG may be predicted by serum proteins. If confirmed in other studies, our findings offer an approach to improved prediction of BAG and enhancing brain health through dietary modifications.

## List of abbreviations

AD: Alzheimer disease
ALKP: alkaline phosphatase
ALT: alanine transaminase
BAG: Brain age gap
BMI: body mass index
BP: Blood pressure
CNN: convolutional neural network
CRP: C-reactive protein
CSF: cerebral-spinal fluid
CVD: cardiovascular disease
DCN: Decorin
ECM: extracellular matrix
FDR: false discovery rate
Gal-9: Galectin 9
green-MED diet: Mediterranean diet higher in polyphenols and lower in red/processed meat
HbA1c: Hemoglobin A1c
HDG: Healthy dietary guidelines
HDLc: High-density lipoprotein cholesterol
HOC: Hippocampal Occupancy Score
IQR: Interquartile Range
LDLc: Low-density lipoprotein cholesterol
LVV: lateral ventricle volume
MCI: Mild cognitive impairment
MED: Mediterranean diet
MRI: Magnetic-Resonance-Imaging
NPX: normalized protein expression
PA: Physical activity
PCA: principal component analysis
PEA: proximity extension assay
RMSE: root mean square error
TG: Triglyceride
WC: Waist circumference

## Acknowledgments

We would like to thank Dr. Assaf Rudich from the Ben-Gurion University of the Negev. We thank the DIRECT PLUS participants for their valuable contributions. We thank MRI technician Eli Atyia and the team from the Nuclear Research Center Negev; Efrat Pupkin, Eyal Goshen, Avi Ben Shabat, and Evyatar Cohen for their valuable contributions to this study.

## Author contribution

AYM, ER, GT, HZ, AK, and IShai conceptualized the DIRECT PLUS and performed the data collection. DP and AYM performed the statistical analysis, interpreted the data, reviewed the literature, and drafted the manuscript. AK performed the MRI-based brain analysis. GL and OF calculate the brain age. IShelef supervised the MRI acquisition. KN performed the proteomic measurements. BI and UC analyzed the data. IShai is the principal investigator of the DIRECT PLUS trial. All authors contributed to the interpretation of data and reviewed this work’s language and intellectual content. MStumvoll, FB, VW, TRR, MB, MSalti, FBH, DDW, MJS, I Shelef, GA, and IShai revised the final draft of the study and approved the final version.

## Ethics approval and consent to participate

The study was conducted in accordance with the Declaration of Helsinki, and the protocol was approved by the Medical Ethics Board and Institutional Review Board at Soroka University Medical Centre, Be’er Sheva, Israel (SOR-0280-16). Participants provided written informed consent and received no compensation.

## Data availability

All data described in the article, code book, and analytic code will be made available upon request, pending the approval of the principal investigator.

