## supplemental File for "Gal-9 and DCN Serum Expression Reflect Accelerated Brain Aging and Are Attenuated by the Green-Mediterranean Diet: The 18-month DIRECT PLUS Proteomics-Brain MRI Trial"

**Supplementary Material:**

**Supplementary Methods 1:**

**Exclusion criteria**

Exclusion criteria were an inability to partake in physical activity, serum creatinine level≥2mg/dL, disturbed liver function, a major illness that might require hospitalization, pregnancy or lactation for femals, presence of active cancer or undergoing chemotherapy either at present or in the prior three years, participation in another trial, chronic treatment with warfarin (given its interaction with vitamin K), and being implanted with a pacemaker or platinum implant.

**Supplementary Methods 2:**

Blood sample analysis

Serum total cholesterol (TC; Coefficient-of-variation (CV), 1.3%), HDL-c, low-density-lipoprotein-cholesterol (LDL-c), and TG (CV, 2.1%) were determined enzymatically with a Cobas-6000 automatic analyzer (Roche). Plasma levels of high-sensitivity C-reactive protein (hsCRP) were measured by ELISA (DiaMed;CV, 1.9%). Plasma glucose levels were measured by Roche GLUC3 (hexokinase method). Plasma insulin levels were measured with an enzyme immunometric assay (Immulite automated analyzer, Diagnostic Products; CV, 2.5%). The homeostatic model of insulin resistance (HOMA IR) was calculated as follows: insulin(μIU/ml)×glucose(mg/dl)/405. All biochemical analyses were performed at the University of Leipzig, Germany.

**Supplementary Methods 3:**

### Certificate of Analysis
### Olink Proteomics

| | |
|---|---|
| **PROJECT NAME** | Klöting_Central_CVD2_17 plates |
| **DELIVERY DATE** | 23.06.2020 |
| **CUSTOMER** | Nora Klöting |
| **ANALYSIS LAB** | Agnese Petrera |
| **QC PERFORMED BY** | Agnese Petrera |
| **REVIEWER** | Katarina Hörnaeus |

http://www.olink.com

#### 1. PROJECT INFORMATION

| Panel name | No. of Samples | No. of Plates | Normalization Method |
|---|---|---|---|
| Olink CARDIOVASCULAR II | 1430 | 17 | Intensity Normalized (v.2) |

##### 1.1 Sample type

human serum

##### 1.2 Project specific comments

There are 20 flagged samples. They deviate in the Internal Controls, and often are timepoints of the same samples, which indicates that the flagging is due to the sample matrix and not a technical error.
1 well empty: plate 11 (F2)
Samples 340-0 (plate 9-G7) and 50-B (plate 11-G5) have been measured in plate 17, as the original measurement failed.

#### 2. QUALITY CONTROL

Four internal controls are added to each sample to monitor the quality of assay performance, as well as the quality of individual samples. The quality control (QC) is performed in two steps:

1. Each sample plate is evaluated on the standard deviation of the internal controls. This should be below 0.2 NPX. Only data from sample plate that pass this quality control will be reported.
2. The quality of each sample is assessed by evaluating the deviation from the median value of the controls for each individual sample. Samples that deviate less than 0.3 NPX from the median pass the quality control.

Data from all samples is included in the data output file. Samples that did not pass the QC are indicated in columns named "QC Warning". Data points from samples that do not pass QC should be treated with caution. [See 4]

##### 2.1 Summary of Quality Control

| Panel name | No. of samples that passed QC / Tot no. of samples | Passed samples (%) |
|---|---|---|
| Olink CARDIOVASCULAR II | 1410 / 1430 | 99 |

##### 2.2 Intra- and Inter-Assay Coefficient of Variance (%CV)

Intra and inter CVs are based on control samples (pooled plasma samples) included on each plate. Calculations are made using linear NPX-values. The number of assays with CVs within defined intervals are presented.

##### 2.2.1 Average %CV

| Panel name | Intra-Assay %CV Reference intra CV <15% | Inter-Assay %CV Reference inter CV <25% |
|---|---|---|
| Olink CARDIOVASCULAR II | 6 | 17 |

##### 2.2.2 Intra-Assay %CV Distribution

| | No. of proteins with %CV within defined intervals | | | | |
|---|---|---|---|---|---|
| Panel name | <5% | 5-10% | 10-15% | >15% | N/A |
| Olink CARDIOVASCULAR II | 36 | 48 | 7 | 1 | 0 |

##### 2.2.3 Inter-Assay %CV Distribution

| Panel name | No. of proteins with %CV within defined intervals <10% | 10-20% | 20-30% | >30% | N/A |
|---|---|---|---|---|---|
| Olink CARDIOVASCULAR II | 16 | 61 | 9 | 6 | 0 |

#### 3. PROTEIN DETECTION RESULTS

##### 3.1 Number of proteins detected in >75% of the samples

| Panel name | No. of detected proteins / Tot no. of proteins | Detected proteins (%) | Expected detectability in EDTA plasma* (%) |
|---|---|---|---|
| Olink CARDIOVASCULAR II | 90 / 92 | 98 | >90 |

*The expected detectability is based on EDTA plasma from healthy donors. These values are intended as guidelines only and protein levels may vary depending on different pathological conditions, sample matrices, or sample preparation methods.

##### 3.2 Data output

Data is presented as normalized protein expression (NPX) values, Olink Proteomics' arbitrary unit on log2 scale. [See 4]
The NPX values are presented in a separate data file. Data points for samples that did not pass QC are written in red text. Data values for measurements below limit of detection (LOD) are reported for all samples. Cells containing data values below LOD are indicated with a pink background. [See 4]

#### 4. FURTHER INFORMATION

Collection of direct links to pages containing important information relating to Olink data generation and processing, as well as additional support content:

https://www.olink.com/key-links/

#### 5. SAMPLES THAT DID NOT PASS QC

| Sample ID | Olink CARDIOVASCULAR II |
|---|---|
| 55-6 | x |
| 88-0 | x |
| 73-6 | x |
| 88-6 | x |
| 73-18 | x |
| 88-18 | x |
| 108-6 | x |
| 108-0 | x |
| 143-0 | x |
| 197-6 | x |
| 295-6 | x |
| 295-18 | x |
| 293-6 | x |
| 295-0 | x |
| 7-A | x |
| 7-B | x |
| 7-C | x |
| 57-A | x |

| Sample ID | Olink CARDIOVASCULAR II |
|---|---|
| 258-A | x |
| 258-C | x |

##### 5.1 Observed deviations

Data for these samples/assays are set to "No data" in the results output file.

| Datapoint | Plate | Reason |
|---|---|---|
| 272-6 / IDUA | NKC_CVD2_plate8 | Datapoint failed |
| 311-0 / IDUA | NKC_CVD2_plate8 | Datapoint failed |

#### Supplementary Methods 4:

#### Packages were used for analyses at R studio:

ggplot2; factoextra; dplyr; tibble; tidyverse; ggpubr; lme4; lmerTest; MASS; quantreg; ppcor; ComplexHeatmap; circlize; gtsummary; broom.

#### Supplementary Figure 1:

#### A flow diagram of the DIRECT PLUS proteomics-brain MRI study

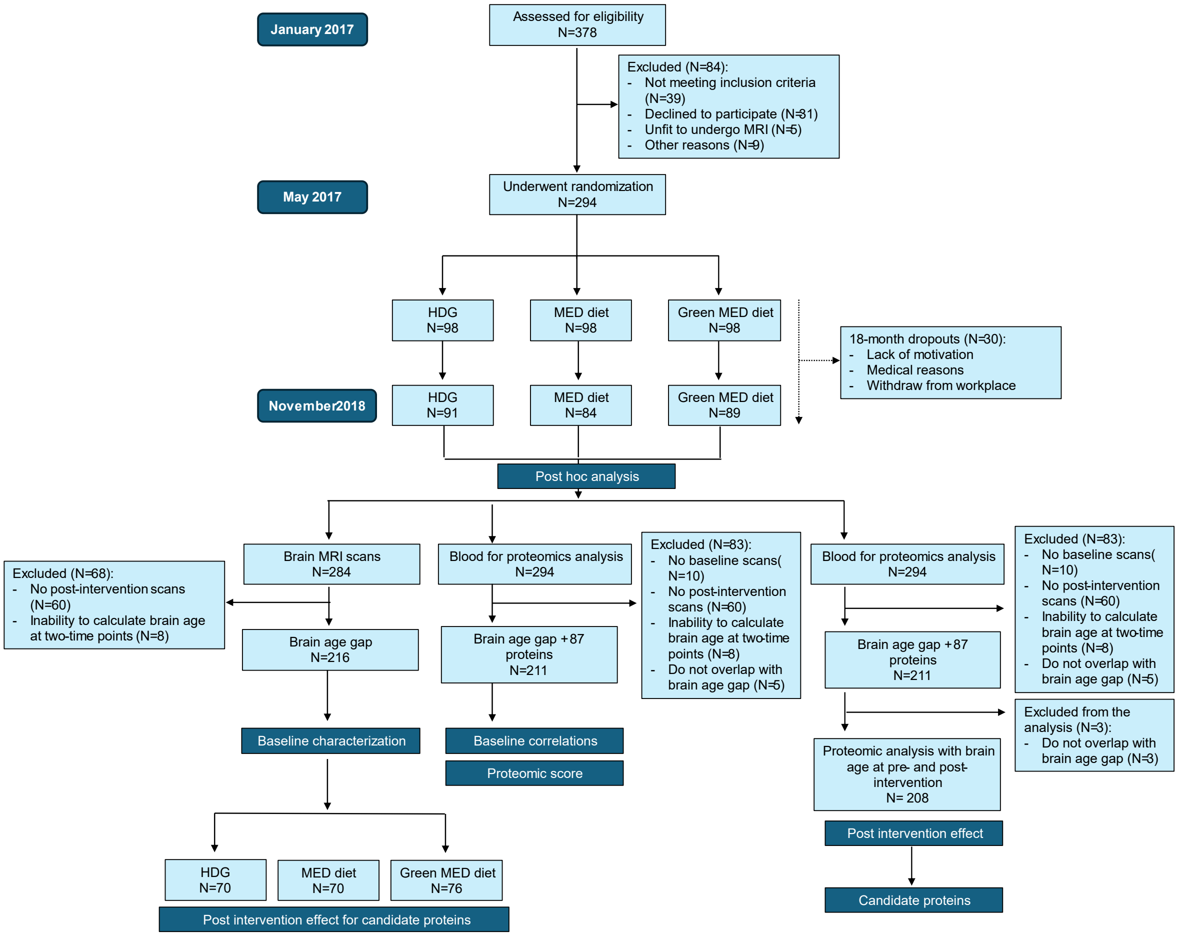

#### Supplementary Figure 2:

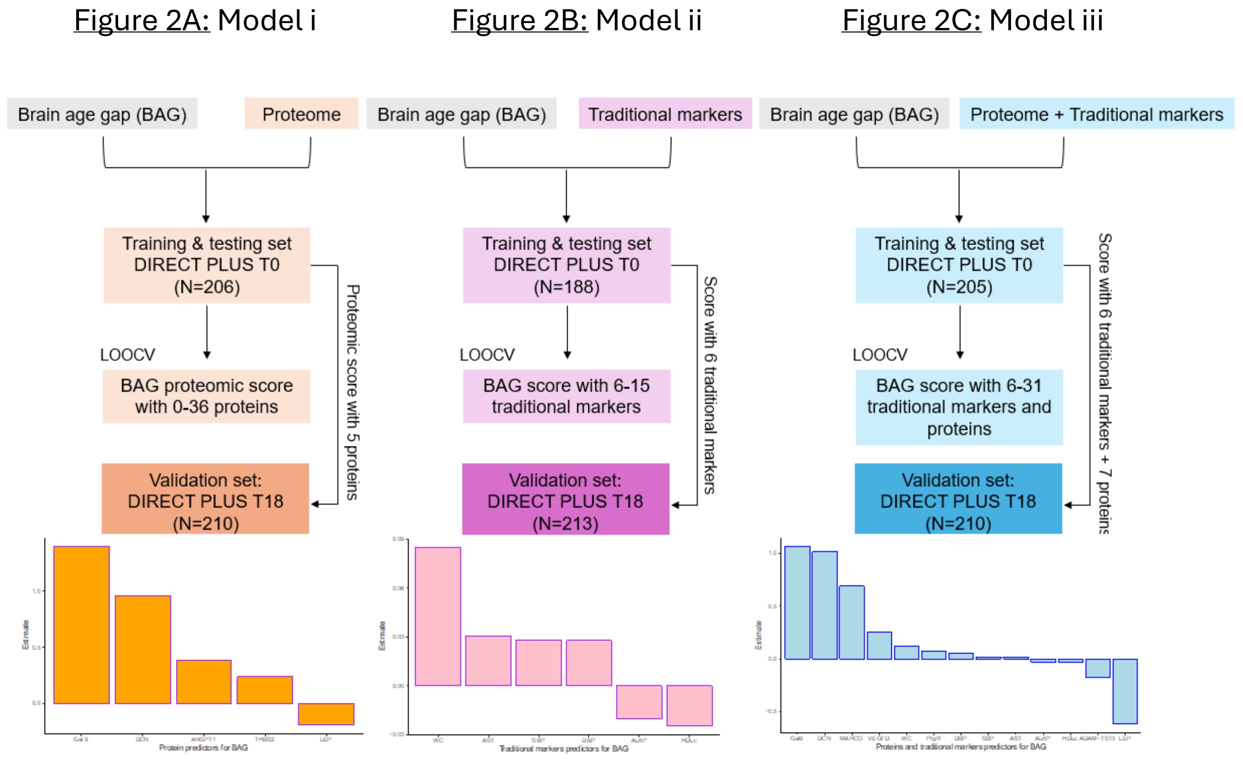


#### Supplementary Table 1:

Prediction of baseline brain age gap (residuals (lm(brain age ~ age))

**Validation of the BAG prediction models**[1]

| Type of models | Sample size for validation | Pearson R |
|---|---|---|
| ***Model i:*** Panel **proteomics** only | 210 | 0.263 (p=1.09e-04) |
| ***Model ii:*** **Traditional markers** only | 213 | 0.275 (p=4.76e-05) |
| ***Model iii:*** Panel **proteomics** with candidate* **traditional markers** | **210** | **0.372 (p=3.52e-08)** |

[1] Validation set included all available T18 observations.

Pearson R was calculated and presented for the validation sets. (i). Baseline panel proteomics to predict brain age gap (residuals (lm (brain age ~ age)) (ii). Baseline traditional markers (see

list below) to predict brain age gap. (iii). Baseline panel proteomics + significant markers from the last fold forced into the model to predict brain age gap.

#### Supplementary Table 2:

Variables with correlations greater than 0.1 in the absolute value of groups 1, 2, and 3 for PC1 component:

| Group 1-Entire[1] | | Group 2-BAG<0[2] | | Group 3-BAG>0[3] | |
|---|---|---|---|---|---|
| **Proteomic Name** | | **Proteomic Name** | | **Proteomic Name** | |
| **ADM** | 0.1 | **ADM** | -0.13 | **ACE2** | 0.11 |
| **AGRP** | 0.1 | **AGRP** | -0.1 | **AMBP** | 0.15 |
| **AMBP** | 0.15 | **AMBP** | -0.15 | **ANGPT1** | 0.11 |
| **ANGPT1** | 0.12 | **ANGPT1** | -0.12 | **CCL3** | 0.12 |
| **BOC** | 0.11 | **BOC** | -0.12 | **CD4** | 0.11 |
| **CCL3** | 0.1 | **CCL17** | -0.1 | **CD40_L** | 0.11 |
| **CD4** | 0.11 | **CD4** | -0.11 | **CD84** | 0.12 |
| **CD40_L** | 0.11 | **CD40_L** | -0.1 | **CEACAM8** | 0.12 |
| **CD84** | 0.13 | **CD84** | -0.14 | **CTSL1** | 0.11 |
| **CEACAM8** | 0.13 | **CEACAM8** | -0.14 | **CXCL1** | 0.13 |
| **CTSL1** | 0.11 | **CTSL1** | -0.1 | **DCN** | 0.12 |
| **CXCL1** | 0.11 | **DCN** | -0.15 | **DECR1** | 0.11 |
| **DCN** | 0.13 | **Dkk_1** | -0.12 | **Dkk_1** | 0.14 |
| **Dkk_1** | 0.13 | **FGF_23** | -0.1 | **Gal_9** | 0.14 |
| **Gal_9** | 0.13 | **Gal_9** | -0.12 | **GDF_2** | 0.11 |
| **GDF_2** | 0.12 | **GDF_2** | -0.12 | **GLO1** | 0.11 |
| **hOSCAR** | 0.14 | **hOSCAR** | -0.14 | **HB_EGF** | 0.12 |
| **HSP_27** | 0.13 | **HSP_27** | -0.11 | **hOSCAR** | 0.13 |
| **IL_1ra** | 0.14 | **IL_1ra** | -0.12 | **HSP_27** | 0.14 |
| **IL_4RA** | 0.11 | **IL_4RA** | -0.11 | **IL_1ra** | 0.15 |
| **IL16** | 0.16 | **IL16** | -0.17 | **IL_4RA** | 0.11 |
| **IL18** | 0.1 | **IL1RL2** | -0.1 | **IL16** | 0.16 |
| **LOX_1** | 0.13 | **LOX_1** | -0.14 | **IL18** | 0.13 |
| **MARCO** | 0.13 | **MARCO** | -0.12 | **IL6** | 0.1 |
| **MERTK** | 0.13 | **MERTK** | -0.15 | **LOX_1** | 0.11 |
| **MMP7** | 0.13 | **MMP7** | -0.14 | **MARCO** | 0.14 |
| **NEMO** | 0.14 | **NEMO** | -0.13 | **MERTK** | 0.11 |
| **PAR_1** | 0.19 | **PAPPA** | -0.11 | **MMP12** | 0.11 |
| **PGF** | 0.15 | **PAR_1** | -0.19 | **MMP7** | 0.13 |
| **PRELP** | 0.13 | **PGF** | -0.16 | **NEMO** | 0.14 |
| **PRSS27** | 0.11 | **PRELP** | -0.16 | **PAR_1** | 0.18 |
| **PRSS8** | 0.14 | **PRSS8** | -0.12 | **PD_L2** | 0.1 |
| **PTX3** | 0.12 | **PTX3** | -0.12 | **PGF** | 0.13 |
| **SOD2** | 0.11 | **SORT1** | -0.16 | **PIgR** | 0.11 |
| **SORT1** | 0.17 | **SPON2** | -0.18 | **PRELP** | 0.11 |
| **SPON2** | 0.18 | **SRC** | -0.11 | **PRSS27** | 0.12 |
| **SRC** | 0.11 | **TF** | -0.11 | **PRSS8** | 0.14 |
| **THBS2** | 0.11 | **THBS2** | -0.11 | **PTX3** | 0.12 |
| **THPO** | 0.1 | **THPO** | -0.12 | **SOD2** | 0.13 |
| **TIE2** | 0.11 | **TIE2** | -0.1 | **SORT1** | 0.17 |
| **TM** | 0.16 | **TM** | -0.16 | **SPON2** | 0.17 |
| **TNFRSF10A** | 0.13 | **TNFRSF10A** | -0.13 | **SRC** | 0.12 |
| **TNFRSF11A** | 0.17 | **TNFRSF11A** | -0.17 | **THBS2** | 0.11 |
| **TNFRSF13B** | 0.1 | **TNFRSF13B** | -0.11 | **TIE2** | 0.1 |
| **TRAIL_R2** | 0.16 | **TRAIL_R2** | -0.16 | **TM** | 0.15 |
| **VSIG2** | 0.11 | **VSIG2** | -0.13 | **TNFRSF10A** | 0.12 |
| | | | | **TNFRSF11A** | 0.16 |

| | |
|---|---|
| **TRAIL_R2** | 0.17 |

[1]Group 1: All participants, n=208. [2]Group 2: Participants who completed with BAG<0, n=102 (Same status- young brain (T0, T18: brain age gap <0)) & (become younger (T0: brain age gap >0, T18: brain age gap <0)). [3]Group 3: Participants who completed with BAG>0, n=106 (Same status- old brain (T0, T18: brain age gap >0)) & (become older (T0: brain age gap <0, T18: brain age gap >0)).

**Supplementary Table 3:**

Significant proteomics list at the volcano plot for each panel group:

| Group 1-Entire[1] | | | Group 2-BAG<0[2] | | | Group 3-bag>0[3] | | |
|---|---|---|---|---|---|---|---|---|
| **Proteomic Name** | **P val** | | **Proteomic Name** | **P val** | | **Proteomic Name** | **P val** | |
| **ADM** | 0 | Positive | **ADM** | 0 | Positive | **ACE2** | 0.209 | No_change |
| **AGRP** | 0.918 | No_change | **AGRP** | 0.776 | No_change | **AMBP** | 0.835 | No_change |
| **AMBP** | 0.358 | No_change | **AMBP** | 0.236 | No_change | **ANGPT1** | 0.745 | No_change |
| **ANGPT1** | 0.294 | No_change | **ANGPT1** | 0.22 | No_change | **CCL3** | 0.029 | Negative |
| **BOC** | 0 | Negative | **BOC** | 0.001 | Negative | **CD4** | 0.04 | Positive |
| **CCL3** | 0.043 | Negative | **CCL17** | 0.609 | No_change | **CD40_L** | 0.001 | Negative |
| **CD4** | 0.005 | Positive | **CD4** | 0.078 | No_change | **CD84** | 0.025 | Negative |
| **CD40_L** | 0 | Negative | **CD40_L** | 0.05 | No_change | **CEACAM8** | 0.015 | Negative |
| **CD84** | 0.016 | Negative | **CD84** | 0.222 | No_change | **CTSL1** | 0.044 | Positive |
| **CEACAM8** | 0.001 | Negative | **CEACAM8** | 0.033 | Negative | **CXCL1** | 0.097 | No_change |
| **CTSL1** | 0.016 | Positive | **CTSL1** | 0.193 | No_change | **DCN** | 0.016 | Positive |
| **CXCL1** | 0.168 | No_change | **DCN** | 0.236 | No_change | **DECR1** | 0.187 | No_change |
| **DCN** | 0.009 | Positive | **Dkk_1** | 0.029 | Negative | **Dkk_1** | 0 | Negative |
| **Dkk_1** | 0 | Negative | **FGF_23** | 0.036 | Negative | **Gal_9** | 0.177 | No_change |
| **Gal_9** | 0.006 | Negative | **Gal_9** | 0.011 | Negative | **GDF_2** | 0.011 | Negative |
| **GDF_2** | 0 | Negative | **GDF_2** | 0.007 | Negative | **GLO1** | 0.186 | No_change |
| **hOSCAR** | 0.003 | Negative | **hOSCAR** | 0.047 | Negative | **HB_EGF** | 0.002 | Negative |
| **HSP_27** | 0 | Negative | **HSP_27** | 0.068 | No_change | **hOSCAR** | 0.029 | Negative |
| **IL_1ra** | 0 | Negative | **IL_1ra** | 0.005 | Negative | **HSP_27** | 0.002 | Negative |
| **IL_4RA** | 0.179 | No_change | **IL_4RA** | 0.275 | No_change | **IL_1ra** | 0.001 | Negative |
| **IL16** | 0.051 | No_change | **IL16** | 0.579 | No_change | **IL_4RA** | 0.404 | No_change |
| **IL18** | 0 | Negative | **IL1RL2** | 0.087 | No_change | **IL16** | 0.032 | Negative |
| **LOX_1** | 0.026 | Negative | **LOX_1** | 0.143 | No_change | **IL18** | 0 | Negative |
| **MARCO** | 0.004 | Negative | **MARCO** | 0.085 | No_change | **IL6** | 0.002 | Negative |
| **MERTK** | 0.399 | No_change | **MERTK** | 0.555 | No_change | **LOX_1** | 0.094 | No_change |
| **MMP7** | 0.091 | No_change | **MMP7** | 0.257 | No_change | **MARCO** | 0.02 | Negative |
| **NEMO** | 0.011 | Negative | **NEMO** | 0.274 | No_change | **MERTK** | 0.556 | No_change |
| **PAR_1** | 0.053 | No_change | **PAPPA** | 0.804 | No_change | **MMP12** | 0.006 | Positive |
| **PGF** | 0.168 | No_change | **PAR_1** | 0.233 | No_change | **MMP7** | 0.247 | No_change |
| **PRELP** | 0.19 | No_change | **PGF** | 0.248 | No_change | **NEMO** | 0.013 | Negative |
| **PRSS27** | 0.002 | Negative | **PRELP** | 0.175 | No_change | **PAR_1** | 0.111 | No_change |
| **PRSS8** | 0 | Negative | **PRSS8** | 0.01 | Negative | **PD_L2** | 0.116 | No_change |
| **PTX3** | 0.806 | No_change | **PTX3** | 0.826 | No_change | **PGF** | 0.348 | No_change |
| **SOD2** | 0.013 | Negative | **SORT1** | 0.027 | Negative | **PIgR** | 0.173 | No_change |
| **SORT1** | 0 | Negative | **SPON2** | 0.072 | No_change | **PRELP** | 0.607 | No_change |
| **SPON2** | 0.008 | Negative | **SRC** | 0.089 | No_change | **PRSS27** | 0.007 | Negative |
| **SRC** | 0 | Negative | **TF** | 0.261 | No_change | **PRSS8** | 0.007 | Negative |
| **THBS2** | 0.711 | No_change | **THBS2** | 0.773 | No_change | **PTX3** | 0.887 | No_change |
| **THPO** | 0.425 | No_change | **THPO** | 0.781 | No_change | **SOD2** | 0.049 | Negative |
| **TIE2** | 0.001 | Negative | **TIE2** | 0.021 | Negative | **SORT1** | 0 | Negative |
| **TM** | 0 | Negative | **TM** | 0.003 | Negative | **SPON2** | 0.052 | No_change |
| **TNFRSF10A** | 0.004 | Negative | **TNFRSF10A** | 0.082 | No_change | **SRC** | 0.001 | Negative |
| **TNFRSF11A** | **0** | Negative | **TNFRSF11A** | 0.01 | Negative | **THBS2** | 0.423 | No_change |
| **TNFRSF13B** | 0.79 | No_change | **TNFRSF13B** | 0.773 | No_change | **TIE2** | 0.019 | Negative |
| **TRAIL_R2** | 0.17 | No_change | **TRAIL_R2** | 0.191 | No_change | **TM** | 0.003 | Negative |

| | | | | | | | | |
|---|---|---|---|---|---|---|---|---|
| **VSIG2** | 0.1 | No_change | **VSIG2** | 0.349 | No_change | **TNFRSF10A** | 0.024 | Negative |
| | | | | | | **TNFRSF11A** | 0.043 | Negative |
| | | | | | | **TRAIL_R2** | 0.381 | No_change |

[1]Group 1: All participants, n=208. [2]Group 2: Participants who completed with BAG<0, n=102 (Same status- young brain (T0, T18: brain age gap <0)) & (become younger (T0: brain age gap >0, T18: brain age gap <0)). [3]Group 3: Participants who completed with BAG>0, n=106 (Same status- old brain (T0, T18: brain age gap >0)) & (become older (T0: brain age gap <0, T18: brain age gap >0)).
